# Impact of stepwise dual antiplatelet therapy de-escalation in patients with multivessel disease undergoing drug-coated balloon angioplasty: insights from the REC-CAGEFREE II trial

**DOI:** 10.64898/2026.08.31.26361869

**Authors:** Yu Zhang, Xingqiang He, Ming Yuan, Fangjun Mou, Jingyu Zhou, Hui Chen, Hua Wang, Wenyi Guo, Yanjin Wei, Zhihui Zhang, Tao Yin, Cheng Zhang, Zhexun Lian, Bin Zhu, Jianzheng Liu, Ruining Zhang, Guotao Fu, Yoshinobu Onuma, Duolao Wang, Patrick W. Serruys, Fu Yi, Chao Gao, Ling Tao

**Author notes:** **Address for correspondence:** Chao Gao, MD, PhD, FESC, Associate Professor of Cardiology, Department of Cardiology, Xijing Hospital, Fourth Military Medical University, Xi’an, 710032, China, The First Affiliated Hospital of USTC, Division of Life Sciences and Medicine, University of Science and Technology of China, Hefei, 230000, China, Fu Yi, MD, PhD, Associate Professor of Cardiology, Department of Cardiology, Xijing Hospital, Fourth Military Medical University, Xi’an 710032, China. These authors contributed equally (Yu Zhang, Xingqiang He, Ming Yuan).

## Abstract

**BACKGROUND:** The optimal antiplatelet regimen in patients with acute coronary syndrome (ACS) and multivessel disease undergoing drug-coated balloon (DCB) angioplasty remains unclear.

**METHODS:** This was a prespecified subgroup analysis of the REC-CAGEFREE II trial, which was conducted at 41 sites in China and randomized 1948 exclusively DCB-treated participants with ACS to stepwise dual antiplatelet therapy (DAPT) de-escalation or standard DAPT. The primary endpoint was net adverse clinical events (NACE; including all-cause death, stroke, myocardial infarction, revascularization, and BARC type 3 or 5 bleeding) at 12 months. Participants were stratified into multivessel and single-vessel subgroups according to angiographic characteristics.

**RESULTS:** Overall, 720/1948 (37.0%) patients had multivessel disease. The multivessel subgroup was associated with a significantly higher risk of NACE compared with the single-vessel subgroup (12.5% versus 6.7%, HR_IPTW_:1.84, 95%CI:1.35-2.51, P<0.001). No significant interaction was observed between vessel status (multivessel or single-vessel) and treatment allocation with respect to NACE (P_interaction_=0.542). In the multivessel subgroup, NACE occurred in 44/368 (12.1%) and 45/352 (12.9%) in the stepwise de-escalation and standard DAPT groups (HR_IPTW_:0.95, 95%CI:0.62-1.75, P=0.818), respectively. In the single-vessel subgroup, NACE occurred in 43/607 (7.1%) and 39/621 (6.3%) in the stepwise de-escalation and standard groups (HR_IPTW_:1.12, 95%CI:0.72-1.70, P=0.611), respectively. For the prespecified hierarchical secondary endpoint, win ratio analyses yielded more wins for stepwise de-escalation in both subgroups.

**CONCLUSIONS:** Among patients with ACS undergoing DCB-only angioplasty, those with multivessel disease were associated with a higher risk of NACE than those with single-vessel disease. Stepwise DAPT de-escalation and standard DAPT exhibited similar risk-benefit profiles in both subgroups.

**Registration Information:** ClinicalTrials.gov identifier: NCT04971356

**CLINICAL PERSPECTIVE:** *What Is New?:*

- This is the first prespecified subgroup analysis that evaluated the efficacy and safety of a stepwise DAPT de-escalation strategy versus standard DAPT among ACS patients with multivessel disease undergoing DCB-only treatment.
- Our findings suggested that stepwise de-escalation was associated with a numerically similar risk of NACE in both multivessel and single-vessel subgroups compared with standard DAPT.

*What Are the Clinical Implications?:*

- For ACS patients undergoing DCB-only angioplasty, stepwise DAPT de-escalation provides similar ischemic protection while reducing bleeding risk irrespective of coronary disease burden. These findings suggest that the stepwise DAPT de-escalation strategy may be considered for this high ischemic risk population.

## INTRODUCTION

Patients presenting with multivessel coronary artery disease usually exhibit more advanced atherosclerotic burden and high ischemic risk, translating into an elevated risk of both in-hospital and long-term recurrent atherothrombotic adverse events.^1–3^ While optimal revascularization strategies for this high-risk population remain a subject of debate, less attention has been paid to the intensity and duration of concomitant antiplatelet regimens. Several meta-analyses have underscored the ischemic benefits of prolonged, intensive dual antiplatelet therapy (DAPT) compared with shorter regimens in patients with multivessel disease undergoing percutaneous coronary intervention (PCI) with drug-eluting stents (DES).^4–6^ However, such prolonged DAPT strategies inherently compromise safety by significantly exacerbating bleeding risks, a concern particularly pronounced in bleeding-vulnerable cohorts. ^4,7^

In contrast to DES, drug-coated balloons (DCBs) obviate the need for the permanent metallic scaffold, thereby facilitating rapid vascular healing free from polymer- or stent-induced chronic inflammation.^8,9^ This favorable biological profile theoretically permits abbreviated or de-escalated antiplatelet strategies after DCB angioplasty.^10^ Recently, the REC-CAGEFREE II trial demonstrated that stepwise DAPT de-escalation (aspirin plus ticagrelor for 1 month, followed by 5 months of ticagrelor monotherapy, and then 6 months of aspirin monotherapy) was non-inferior to the standard 12-month DAPT (aspirin plus ticagrelor) with respect to 12-month net adverse clinical events (NACE) in acute coronary syndrome (ACS) patients undergoing exclusive DCB angioplasty.^11^ Previous studies evaluating antiplatelet strategies after PCI have mainly focused on the DES scenario,^12–17^ whereas evidence regarding the optimal antiplatelet regimen in ACS patients with multivessel disease undergoing DCB angioplasty remains uncertain.

In this prespecified subgroup analysis from the REC-CAGEFREE II trial, we aimed to compare the efficacy and safety of stepwise DAPT de-escalation versus standard 12-month DAPT among ACS patients with multivessel or single-vessel disease undergoing DCB-only angioplasty.

## METHODS

### Study Design and Population

The REC-CAGEFREE II was an investigator-initiated, multicenter, randomized, open-label, non-inferiority trial conducted at 41 sites in China from November 27, 2021, to January 21, 2023. The trial randomized 1948 patients with ACS, all treated exclusively with paclitaxel-coated balloons, to stepwise DAPT de-escalation (1 month of aspirin plus ticagrelor, followed by 5 months of ticagrelor monotherapy and 6 months of aspirin monotherapy) or standard DAPT (12 months of aspirin plus ticagrelor) (ClinicalTrials.gov identifier: NCT04971356). The rationale, study design, inclusion/exclusion criteria, and primary results of the trial have been previously published.^11,18^ The institutional review board at each participating site approved the trial protocol. The study complied with the Declaration of Helsinki and Good Clinical Practice. All participants provided written informed consent.

In the REC-CAGEFREE II trial, all participants with a clinical presentation of ACS (ST/non-ST elevation myocardial infarction (MI) or unstable angina) and undergoing successful DCB-only treatment were eligible for enrollment. The selection of suitable patients/lesions for DCB treatment and subsequent procedural techniques followed the recommendations of the German Consensus Group on DCBs for treatment of coronary artery disease^19^ and the Third Report of the International DCB Consensus Group.^9^ No restrictions were imposed regarding lesion type (*de novo* or in-stent restenosis), treated vessel diameter, or the specific brand of paclitaxel-coated balloon used. Key exclusion criteria included age below 18 or above 80 years, prior hemorrhagic stroke, requirement for long-term oral anticoagulation, cardiogenic shock, and presentation with in-stent thrombosis. Immediately after DCB angioplasty, patients were randomly assigned in a 1:1 ratio via a web-based centralized system to receive either stepwise DAPT de-escalation or standard 12-month DAPT.

In this prespecified subgroup analysis, participants were stratified into multivessel and single-vessel subgroups according to coronary angiographic characteristics. Multivessel coronary artery disease was defined as a diameter stenosis greater than 50% by visual assessment in more than one major coronary vessel at the time of the index PCI (with or without prior revascularization).

### Clinical Outcomes and Follow-up

The primary efficacy endpoint was NACE, a non-hierarchical composite endpoint comprising all-cause death, stroke, myocardial infarction (MI), revascularization, and Bleeding Academic Research Consortium (BARC) type 3 or 5 bleeding, evaluated at 12-month follow-up. The secondary efficacy endpoints included a hierarchical composite of clinically relevant ischemic or bleeding events, BARC type 2, 3, or 5 bleeding, and BARC type 3 or 5 bleeding. A clinically relevant ischemic or bleeding event was predefined. This endpoint was analyzed with the win ratio method in the following order of clinical priority: all-cause death, stroke, MI, BARC type 3 bleeding, revascularization, and BARC type 2 bleeding.^20^ The safety endpoints included the patient-oriented composite endpoint (PoCE, defined as a non-hierarchical composite of all-cause death, stroke, MI, and revascularization), device-oriented composite endpoint (DoCE, defined as a non-hierarchical composite of cardiovascular death, target vessel myocardial infarction, and clinically and physiologically indicated target lesion revascularization), and their individual components. All endpoint events were adjudicated by an independent clinical event committee, in accordance with the definitions from the Academic Research Consortium (ARC)-2,^21^ the Fourth Universal Definition of MI for spontaneous MI,^22^ and BARC.^23^ Adverse events were collected centrally, and any document that could lead to unblinding of treatment-group assignment was redacted before submission to the clinical event committee.

Scheduled follow-up visits occurred at 1 (±14 days), 3, 6, and 12 (±30 days) months after randomization. Visits were preferably conducted on-site; however, if patients were unable or unwilling to visit the outpatient clinic, the scheduled visit could be replaced by a telephone call, except for the 30-day and 12-month visits.

### Statistical Analysis

The primary analyses were conducted in the intention-to-treat (ITT) population, defined as all patients who provided informed consent and were randomly assigned to treatment. Outcomes of patients who were lost to follow-up or withdrew consent were censored at their date of last contact. Continuous variables are summarized as means ± standard deviations, depending on their distribution. Categorical variables are expressed as numbers (percentages). Continuous variables were compared with the Student’s t-test or the Wilcoxon rank-sum test based on the data distribution. Categorical variables were compared using the Chi-square test or Fisher’s exact test. The 12-month cumulative incidences of primary and secondary endpoints were estimated using the Kaplan-Meier method. Cox proportional hazards regression models were used to estimate hazard ratios (HRs) alongside 95% confidence intervals (CIs). When no events were observed in a treatment group, HRs and 95% CIs were estimated using Cox proportional hazards models with Firth’s penalized partial likelihood correction.^24^ The secondary endpoint of clinically relevant ischemic or bleeding events was analyzed using the unmatched win ratio method,^25^ which sequentially compared patient pairs between groups based on the prespecified hierarchy of events. Treatment effect heterogeneity across subgroups was evaluated using interaction terms in Cox models for time-to-event endpoints and Z-tests of log-transformed win-ratio estimates.^20^ Inverse probability of treatment weighting (IPTW) and multivariable Cox regression were applied to account for potential baseline imbalances between groups. IPTW weights were estimated via logistic regression, including the covariates of age, sex, smoking, hypertension, diabetes mellitus, hyperlipidemia, left ventricular ejection fraction, previous MI, previous PCI, previous lacunar infarction, chronic obstructive pulmonary disease (COPD), chronic kidney disease (CKD), estimated glomerular filtration rate (eGFR), clinical presentation, SYNTAX score, GRACE score, and PRECISE-DAPT score. Multivariable Cox models included age, sex, smoking status, hypertension, diabetes mellitus, hyperlipidemia, prior lacunar infarction, COPD, and CKD. To assess result robustness, sensitivity analyses were conducted within the per-protocol (PP) population. All analyses were performed with R version 4.4.3 (R Foundation for Statistical Computing, Vienna, Austria). Statistical significance was defined as a two-tailed P-value, with P < 0.05 representing the significance threshold.

## RESULTS

### Baseline Characteristics

Between November 27, 2021, and January 21, 2023, a total of 1948 eligible participants were enrolled in the REC-CAGEFREE II trial. Of these, 720/1948 (37.0%) had multivessel disease and 1228/1948 (63.0%) had single-vessel disease (Figure 1). Compared with patients in the single-vessel subgroup, those with multivessel disease were older, more likely to be male, and had a higher prevalence of comorbidities, including hyperlipidemia, hypertension, diabetes mellitus, peripheral arterial disease, previous MI, previous PCI, and CKD. In terms of procedural and lesion characteristics, patients in the multivessel subgroup were less likely to undergo radial-access PCI, achieve complete revascularization, and were treated with more and longer DCBs, compared with those in the single-vessel subgroup. The baseline characteristics of patients receiving stepwise de-escalation versus standard DAPT were generally well balanced in the two subgroups (Table 1). Medication adherence rates and reasons for nonadherence across both subgroups were presented in Table S1 and Figure S1.

**Figure 1.**
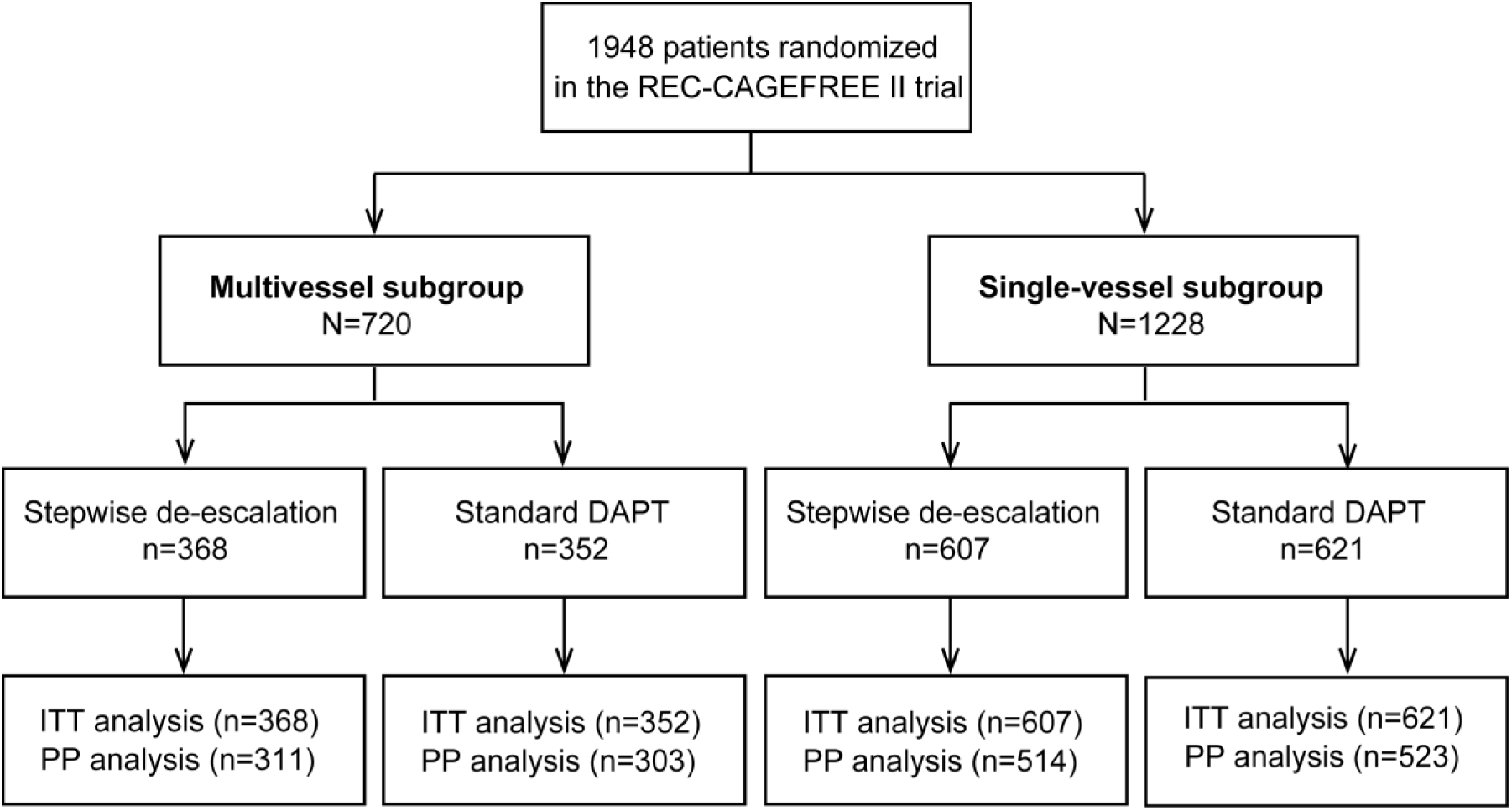
Study flowchart. DAPT, dual antiplatelet therapy; ITT, intention-to-treat; PP, per-protocol.

**Table 1.** Baseline characteristics.

| Baseline characteristics | Multivessel<br>(N = 720) | Single-vessel<br>(N = 1228) | P | Multivessel (N = 720) |  |  | Single-vessel (N = 1228) |  |  |
| --- | --- | --- | --- | --- | --- | --- | --- | --- | --- |
|  |  |  |  | Stepwise<br>de-escalation<br>(n = 368) | Standard<br>DAPT<br>(n = 352) | P | Stepwise<br>de-escalation<br>(n = 607) | Standard<br>DAPT<br>(n = 621) | P |
| Demographic characteristics |  |  |  |  |  |  |  |  |  |
| Age, years | 60.0 ± 10.6 | 58.7 ± 10.9 | 0.011 | 59.9 ± 10.4 | 60.2 ± 10.8 | 0.722 | 59.1 ± 10.8 | 58.3 ± 11.1 | 0.221 |
| Male, n (%) | 567 (78.8) | 893 (72.7) | 0.003 | 288 (78.3) | 279 (79.3) | 0.743 | 439 (72.3) | 454 (73.1) | 0.758 |
| BMI, kg/m<sup>2</sup> | 25.1 ± 3.2 | 25.1 ± 3.5 | 0.668 | 25.1 ± 3.1 | 25.1 ± 3.2 | 0.983 | 24.9 ± 3.4 | 25.2 ± 3.5 | 0.099 |
| Current smoker, n (%) | 254 (36.4) | 415 (34.3) | 0.338 | 120 (33.6) | 134 (39.4) | 0.112 | 188 (31.5) | 227 (37.0) | 0.052 |
| Hyperlipidemia, n (%) | 594 (84.5) | 591 (78.8) | 0.003 | 300 (83.1) | 294 (86.0) | 0.345 | 461 (77.3) | 490 (80.2) | 0.255 |
| Left ventricular ejection fraction, % | 58.3 ± 8.3 | 60.1 ± 8.0 | <0.001 | 58.4 ± 8.3 | 58.6 ± 8.4 | 0.763 | 60.2 ± 8.3 | 60.1 ± 7.6 | 0.765 |
| Hypertension, n (%) | 474 (65.8) | 703 (57.2) | <0.001 | 236 (64.1) | 238 (67.6) | 0.365 | 347 (57.2) | 356 (57.3) | >0.999 |
| Diabetes mellitus, n (%) | 267 (37.1) | 328 (26.7) | <0.001 | 135 (36.7) | 132 (37.5) | 0.881 | 153 (25.2) | 175 (28.2) | 0.266 |
| Insulin-treated, n (%) | 63 (25.0) | 88 (28.4) | 0.510 | 34 (26.4) | 29 (23.6) | 0.251 | 37 (26.6) | 51 (29.8) | 0.514 |
| Chronic kidney disease, n (%) | 57 (7.9) | 60 (4.9) | 0.009 | 24 (6.5) | 33 (9.4) | 0.201 | 34 (5.6) | 26 (4.2) | 0.309 |
| Peripheral arterial disease, n (%) | 28 (3.9) | 27 (2.2) | 0.042 | 15 (4.1) | 13 (3.7%) | 0.948 | 16 (2.6) | 11 (1.8) | 0.402 |
| Previous MI, n (%) | 135 (18.8) | 121 (9.9) | <0.001 | 66 (18.0) | 69 (19.6) | 0.645 | 69 (11.4) | 52 (8.4) | 0.098 |
| Previous PCI, n (%) | 276 (38.3) | 351 (28.6) | <0.001 | 133 (36.1) | 143 (40.6) | 0.246 | 186 (30.6) | 165 (26.6) | 0.130 |
| Previous CABG, n (%) | 7 (1.0) | 3 (28.6) | 0.066 | 4 (1.1) | 3 (0.9) | >0.999 | 1 (0.2) | 2 (0.3) | >0.999 |
| Previous stroke, n (%) | 75 (10.4) | 96 (7.8) | 0.062 | 43 (11.7) | 32 (9.1) | 0.315 | 50 (8.3) | 46 (7.4) | 0.657 |
| COPD, n (%) | 24 (3.5) | 49 (4.2) | 0.556 | 18 (5.0) | 6 (1.8) | 0.037 | 25 (4.3) | 24 (4.0) | 0.908 |
| High bleeding risk*, n (%) | 157 (22.6) | 230 (19.5) | 0.123 | 79 (22.4) | 78 (22.7) | 0.998 | 118 (20.2) | 112 (18.8) | 0.590 |
| Clinical presentation, n (%) |  |  | 0.002 |  |  | 0.554 |  |  | 0.952 |
| ST-elevation MI | 126 (17.5) | 200 (16.3) |  | 59 (16.0) | 67 (19.0) |  | 100 (16.5) | 100 (16.1) |  |
| Non-ST-elevation MI | 226 (31.4) | 306 (24.9) |  | 119 (32.3) | 107 (30.4) |  | 149 (24.5) | 157 (25.3) |  |
| Unstable angina | 368 (51.1) | 722 (58.8) |  | 190 (51.6) | 178 (50.6) |  | 358 (59.0) | 364 (58.6) |  |
| SYNTAX score | 8.0 ± 5.9 | 6.0 ± 4.2 | <0.001 | 7.6 ± 5.2 | 8.5 ± 6.5 | 0.049 | 5.8 ± 4.0 | 6.1 ± 4.3 | 0.152 |
| GRACE score | 87.4 ± 23.9 | 84.2 ± 23.3 | 0.004 | 86.5 ± 24.5 | 88.2 ± 23.4 | 0.349 | 85.2 ± 23.9 | 83.2 ± 22.6 | 0.146 |
| PRECISE-DAPT score, n (%) |  |  | 0.007 |  |  | 0.233 |  |  | 0.642 |
| 0-25 | 644 (93.9) | 1111 (95.5) |  | 332 (95.4) | 312 (92.3) |  | 546 (95.0) | 565 (96.1) |  |
| ≥25 | 42 (6.1) | 52 (4.5) |  | 16 (4.6) | 26 (7.7) |  | 29 (5.0) | 23 (3.9) |  |
| **Procedural characteristics** |  |  |  |  |  |  |  |  |  |
| Radial access approach, n (%) | 666 (92.5) | 1173 (95.5) | 0.011 | 345 (93.8) | 321 (91.2) | 0.222 | 582 (95.9) | 591 (95.2) | 0.772 |
| IVUS/OCT, n (%) | 82 (11.4) | 170 (13.8) | 0.137 | 38 (10.3) | 44 (12.5) | 0.423 | 90 (14.8) | 80 (12.9) | 0.366 |
| Complete revascularization, n (%) | 510 (72.5) | 1093 (91.8) | <0.001 | 273 (75.8) | 237 (69.1) | 0.055 | 539 (90.7) | 554 (92.8) | 0.236 |
| Number of DCBs used per patient | 1.4 ± 0.9 | 1.1 ± 0.4 | <0.001 | 1.6 ± 0.9 | 1.6 ± 0.9 | 0.491 | 1.1 ± 0.4 | 1.1 ± 0.4 | 0.890 |
| Total DCB length, mm | 41.1 ± 26.4 | 27.8 ± 12.8 | <0.001 | 41.6 ± 26.4 | 40.5 ± 26.4 | 0.593 | 27.4 ± 12.4 | 28.2 ± 13.3 | 0.290 |
| Mean diameter of DCB, mm | 2.6 ± 0.4 | 2.8 ± 0.5 | <0.001 | 2.6 ± 0.4 | 2.6 ± 0.4 | 0.367 | 2.8 ± 0.5 | 2.8 ± 0.5 | 0.946 |
| Lesion location, n (%) |  |  | <0.001 |  |  | 0.955 |  |  | 0.646 |
| Left main | 5 (0.5) | 3 (0.2) |  | 3 (0.6) | 2 (0.4) |  | 1 (0.2) | 2 (0.3) |  |
| Left anterior descending artery | 344 (34.3) | 738 (56.0) |  | 177 (34.4) | 167 (34.2) |  | 378 (57.6) | 360 (54.5) |  |
| Left circumflex artery | 356 (35.5) | 340 (25.8) |  | 185 (35.9) | 171 (35.0) |  | 162 (24.7) | 178 (26.9) |  |
| Right coronary artery | 298 (29.7) | 236 (17.9) |  | 150 (29.1) | 148 (30.3) |  | 115 (17.5) | 121 (18.3) |  |
| Vessel disease, n (%) |  |  | <0.001 |  |  | 0.855 |  |  | - |
| 1-VD | 0 (0) | 1228 (100.0) |  | 0 (0) | 0 (0) |  | 607 (100.0) | 621 (100.0) |  |
| 2-VD | 533 (74.0) | 0 (0) |  | 274 (74.5) | 259 (73.6) |  | 0 (0) | 0 (0) |  |
| 3-VD | 187 (26.0) | 0 (0) |  | 94 (25.5) | 93 (26.4) |  | 0 (0) | 0 (0) |  |
| **Target lesion characteristics** |  |  |  |  |  |  |  |  |  |
| In-stent restenosis, n (%) | 192 (19.1) | 222 (16.9) | 0.171 | 90 (17.5) | 102 (20.9) | 0.194 | 118 (18.0) | 104 (15.7) | 0.308 |
| Small vessel (<3.0 mm), n (%) | 666 (66.4) | 748 (56.8) | <0.001 | 346 (67.2) | 320 (65.6) | 0.636 | 371 (56.6) | 377 (57.0) | 0.904 |
| Bifurcation<sup>†</sup>, n (%) | 382 (39.0) | 583 (45.5) | 0.002 | 207 (41.2) | 175 (36.8) | 0.180 | 288 (45.0) | 295 (46.1) | 0.736 |
| Long lesion (≥28 mm), n (%) | 448 (44.7) | 430 (32.6) | <0.001 | 234 (45.4) | 214 (43.9) | 0.659 | 195 (29.7) | 235 (35.6) | 0.028 |

Data are presented based on the intention-to-treat population. Values are expressed as mean ± SD for continuous variables and n (%) for categorical variables. BMI, body mass index;
CABG, coronary artery bypass graft; COPD, chronic obstructive pulmonary disease; DAPT, dual antiplatelet therapy; DCB, drug-coated balloon; IVUS, intravascular ultrasound;
MI, myocardial infarction; OCT, optical coherence tomography; PCI, percutaneous coronary intervention. \*Defined according to the Academic Research Consortium for High
Bleeding Risk. <sup>†</sup>Bifurcation was classified when at least 50% luminal diameter stenosis occurred within 3 mm of the bifurcation point, according to the SYNTAX score definition.

### Clinical Outcomes in the Multivessel and Single-Vessel Subgroups

In the ITT population, the primary endpoint NACE occurred in 89 (12.5%) of 720 patients in the multivessel subgroup and 82 (6.7%) of 1228 patients in the single-vessel subgroup at 12-month follow-up (HR_IPTW_:1.84, 95% CI: 1.35-2.51, P < 0.001; Figure 2A and Table S2). Win ratio analysis demonstrated significantly fewer wins in the multivessel subgroup compared with the single-vessel subgroup (10.4% vs 15.1%; win ratio_IPTW_: 0.73, 95% CI 0.56-0.94, P = 0.015; Table S2). The results of other secondary endpoints are shown in Figure 2B-D and Table S2.

**Figure 2.**
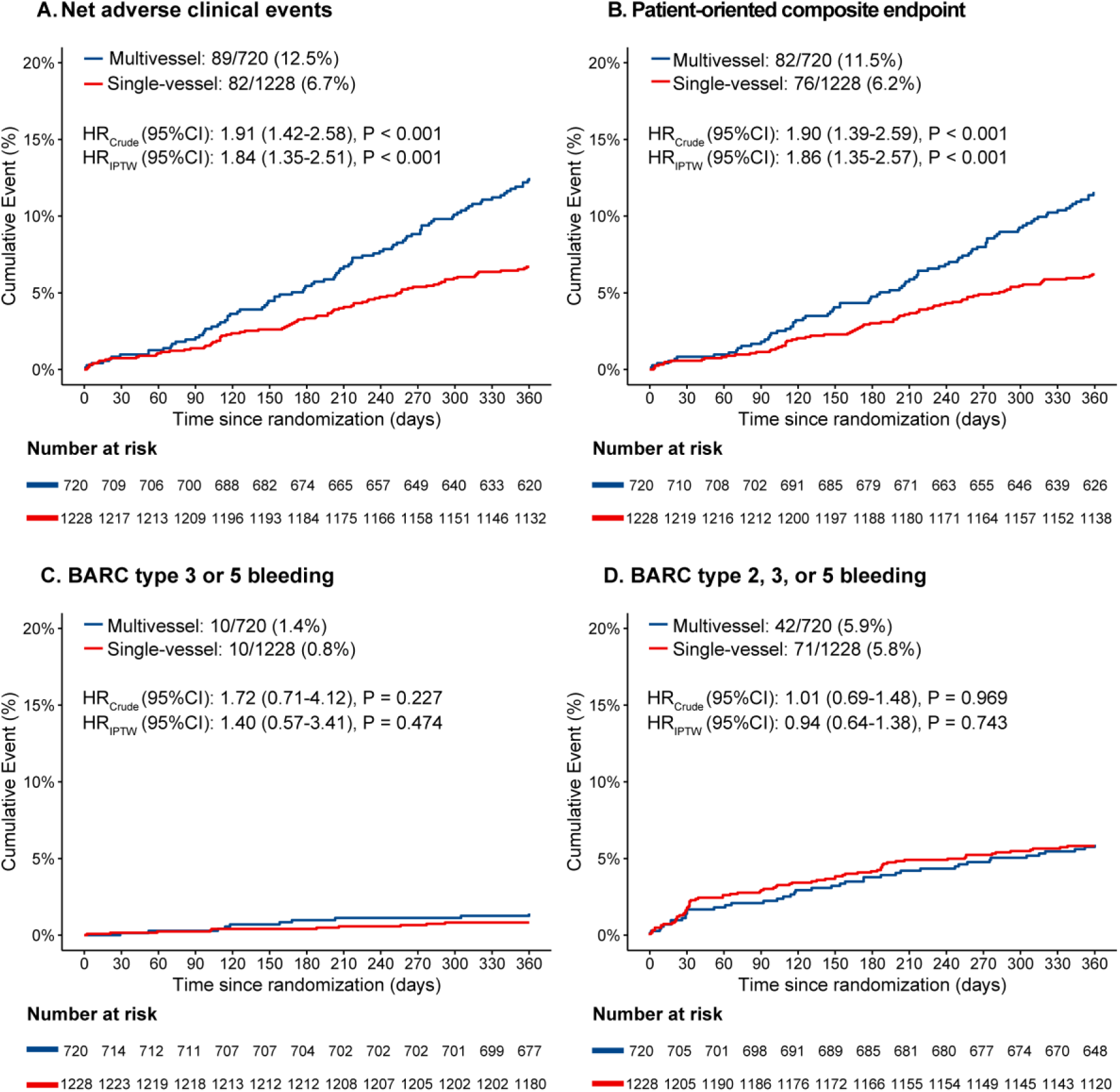
Kaplan-Meier curves of primary and secondary endpoints stratified by subgroups. **A**, Net adverse clinical event; **B**, Patient-oriented composite endpoint; **C**, BARC type 3 or 5 bleeding; **D**, BARC type 2, 3, or 5 bleeding. BARC, Bleeding Academic Research Consortium; CI, confidence interval; HR, hazard ratio; IPTW, inverse probability of treatment weighting.

### Clinical Outcomes by Vessel Disease Subgroups and Treatment Allocation

No significant interaction was observed between vessel disease subgroups (multivessel versus single-vessel) and treatment allocation for the NACE (P_interaction_ = 0.542). In the multivessel subgroup, NACE occurred in 44 (12.1%) of 368 patients in the stepwise de-escalation group versus 45 (12.9%) of 352 patients in the standard DAPT group (HR_IPTW_: 0.95, 95% CI: 0.62-1.75, P = 0.818). In the single-vessel subgroup, the corresponding NACE rates were 43/607 (7.1%) and 39/621 (6.3%), respectively (HR_IPTW_: 1.12, 95% CI: 0.72-1.75, P = 0.611; Figure 3A and Table 2). The results of individual components of NACE are shown in Figure S2 and Table S4. For bleeding outcomes, the stepwise de-escalation was associated with a significantly lower risk of BARC type 2, 3, or 5 bleeding compared with standard DAPT in both subgroups (multivessel subgroup: 2.8% versus 9.2%, HR_IPTW_: 0.29, 95% CI: 0.14-0.59, P = 0.002; single-vessel subgroup: 2.2% versus 9.4%, HR_IPTW_: 0.21, 95% CI: 0.12-0.40, P < 0.001; P_interaction_ = 0.564; Figure 3D). The results of other secondary endpoints are summarized in Figure 3B-C and Table 2.

**Figure 3.**
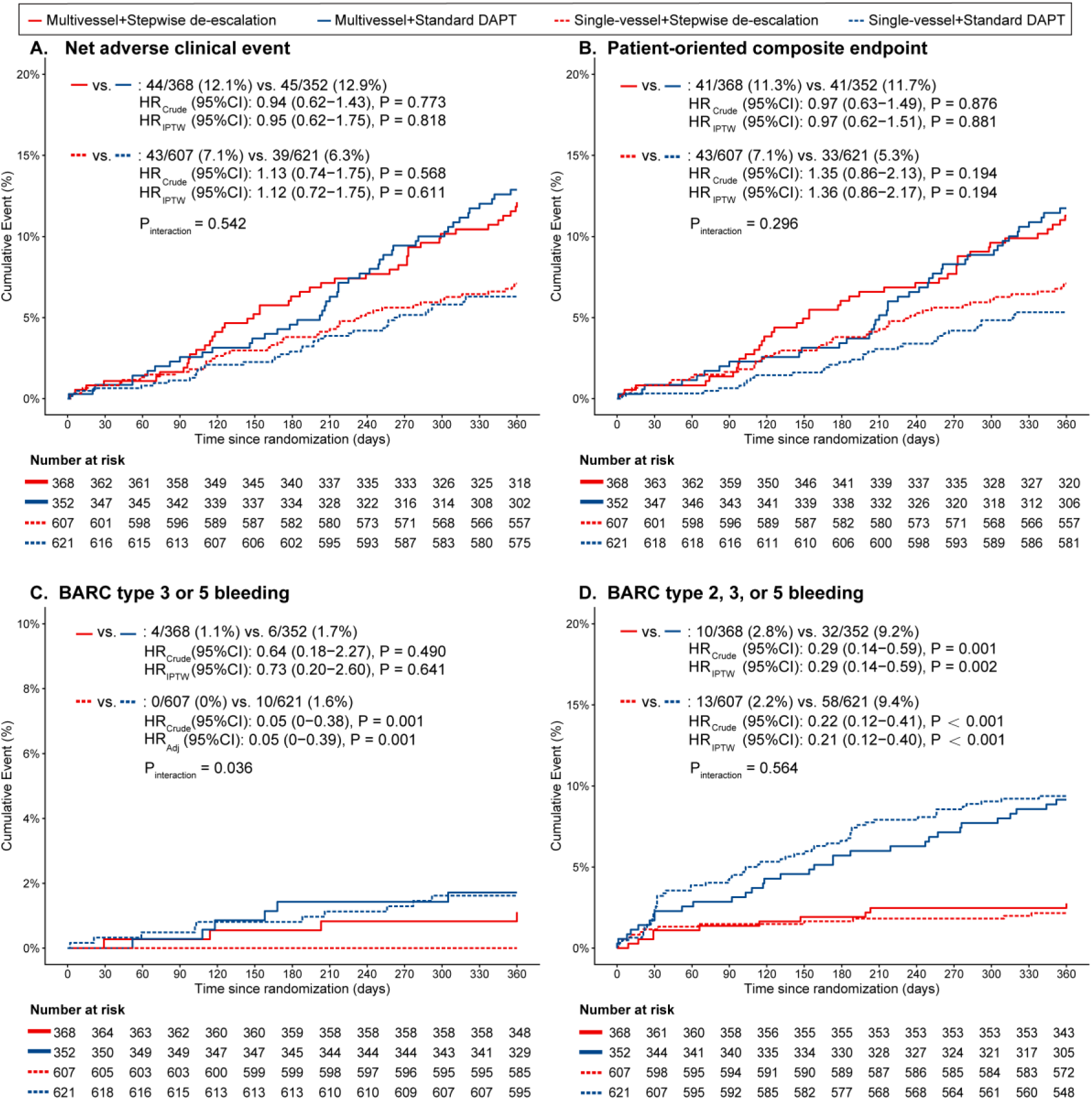
Kaplan-Meier curves of primary and secondary endpoints stratified by subgroups and treatment allocation. **A**, Net adverse clinical event; **B**, Patient-oriented composite endpoint; **C**, BARC type 3 or 5 bleeding; **D**, BARC type 2, 3, or 5 bleeding. For **A**, **B**, and **D**, IPTW was applied to adjust for confounding. For **C**, Firth’s penalized Cox regression was used to account for zero events observed in the stepwise de-escalated group among patients with single-vessel disease. BARC, Bleeding Academic Research Consortium; CI, confidence interval; DAPT, dual antiplatelet therapy; HR, hazard ratio; IPTW, inverse probability of treatment weighting.

**Table 2.** Clinical outcomes at 12 months stratified by subgroups and treatment allocation.

| Outcomes | Multivessel (N = 720) |  |  |  |  | Single-vessel (N = 1228) |  |  |  |  | P for interaction |
| --- | --- | --- | --- | --- | --- | --- | --- | --- | --- | --- | --- |
|  | Stepwise de-escalation (n = 368) | Standard DAPT (n = 352) | Adjusted method | HR (95% CI) | P | Stepwise de-escalation (n = 607) | Standard DAPT (n = 621) | Adjusted method | HR (95% CI) | P |  |
| Primary endpoint |  |  |  |  |  |  |  |  |  |  |  |
| Net adverse clinical events | 44 (12.1) | 45 (12.9) | Crude<br>IPTW | 0.94 (0.62-1.43)<br>0.95 (0.62-1.75) | 0.773<br>0.818 | 43 (7.1) | 39 (6.3) | Crude<br>IPTW | 1.13 (0.74-1.75)<br>1.12 (0.72-1.75) | 0.568<br>0.611 | 0.542 |
| Secondary endpoints |  |  |  |  |  |  |  |  |  |  |  |
| Clinically relevant ischemic or bleeding event* | 22417 (17.3) | 15551 (12.0) | Win ratio<br>IPTW | 1.44 (1.00-2.08)<br>1.48 (1.02-2.16) | 0.051<br>0.041 | 48152 (12.8) | 33230 (8.8) | Win ratio<br>IPTW | 1.45 (1.04-2.02)<br>1.47 (1.05-2.07) | 0.028<br>0.025 | 0.984 |
| BARC type 3 or 5 bleeding | 4 (1.1) | 6 (1.7) | Crude<br>IPTW | 0.64 (0.18-2.27)<br>0.73 (0.20-2.60) | 0.490<br>0.641 | 0 (0) | 10 (1.6) | Crude<br>IPTW | 0.05 (0-0.38)<br>- | 0.001<br>- | 0.036 |
| BARC type 2, 3, or 5 bleeding | 10 (2.8) | 32 (9.2) | Crude<br>IPTW | 0.29 (0.14-0.59)<br>0.29 (0.14-0.59) | 0.001<br>0.002 | 13 (2.2) | 58 (9.4) | Crude<br>IPTW | 0.22 (0.12-0.41)<br>0.21 (0.12-0.40) | <0.001<br><0.001 | 0.564 |
| BARC type 2 bleeding | 6 (1.7) | 27 (7.7) | Crude<br>IPTW | 0.21 (0.09-0.51)<br>0.20 (0.08-0.50) | 0.001<br>0.002 | 13(2.2) | 48 (7.8) | Crude<br>IPTW | 0.27 (0.15-0.50)<br>0.26 (0.14-0.49) | <0.001<br><0.001 | 0.636 |
| Device-oriented composite endpoint | 21 (5.8) | 21 (6.0) | Crude<br>IPTW | 0.97 (0.53-1.78)<br>0.96 (0.51-1.80) | 0.923<br>0.901 | 30 (5.0) | 24 (3.9) | Crude<br>IPTW | 1.29 (0.76-2.21)<br>1.26 (0.73-2.17) | 0.350<br>0.420 | 0.489 |
| Cardiovascular death | 3 (0.8) | 2 (0.6) | Crude<br>IPTW | 1.45 (0.24-8.65)<br>1.41 (0.23-8.58) | 0.686<br>0.735 | 10 (1.7) | 4 (0.6) | Crude<br>IPTW | 2.57 (0.81-8.20)<br>2.20 (0.67-7.16) | 0.110<br>0.218 | 0.598 |
| Target vessel myocardial infarction | 2 (0.6) | 3 (0.9) | Crude<br>IPTW | 0.64 (0.11-3.85)<br>0.63 (0.10-4.18) | 0.629<br>0.672 | 5 (0.8) | 5 (0.8) | Crude<br>IPTW | 1.03 (0.30-3.56)<br>1.09 (0.31-3.82) | 0.960<br>0.901 | 0.670 |
| Clinically and physiologically indicated lesion revascularization | 18 (5.0) | 17 (4.9) | Crude<br>IPTW | 1.03 (0.53-2.00)<br>1.04 (0.52-2.07) | 0.926<br>0.912 | 19 (3.2) | 18 (2.9) | Crude<br>IPTW | 1.09 (0.57-2.08)<br>1.13 (0.58-2.18) | 0.789<br>0.723 | 0.905 |
| Patient-oriented composite endpoint | 41 (11.3) | 41 (11.7) | Crude<br>IPTW | 0.97 (0.63-1.49)<br>0.97 (0.62-1.51) | 0.876<br>0.881 | 43 (7.1) | 33 (5.3) | Crude<br>IPTW | 1.35 (0.86-2.13)<br>1.36 (0.86-2.17) | 0.194<br>0.194 | 0.296 |
| All-cause death | 3 (0.8) | 3 (0.9) | Crude<br>IPTW | 0.97 (0.19-4.78)<br>0.94 (0.19-4.71) | 0.965<br>0.940 | 10 (1.7) | 4 (0.6) | Crude<br>IPTW | 2.57 (0.81-8.20)<br>2.20 (0.67-7.16) | 0.110<br>0.218 | 0.331 |
| Stroke | 3 (0.8) | 3 (0.9) | Crude | 0.97 (0.19-4.79) | 0.966 | 4 (0.7) | 5 (0.8) | Crude | 0.82 (0.22-3.07) | 0.772 | 0.880 |
|  |  |  | IPTW | 1.16 (0.23-5.77) | 0.867 |  |  | IPTW | 0.79 (0.21-3.02) | 0.741 |  |
| Myocardial infarction | 2 (0.6) | 5 (1.4) | Crude | 0.38 (0.07-1.98) | 0.253 | 7 (1.2) | 5 (0.8) | Crude | 1.45 (0.46-4.56) | 0.585 | 0.194 |
|  |  |  | IPTW | 0.37 (0.07-2.10) | 0.321 |  |  | IPTW | 1.67 (0.52-5.41) | 0.412 |  |
| Revascularization | 36 (10.0) | 35 (10.0) | Crude | 1.00 (0.63-1.59) | 0.993 | 29 (4.8) | 26 (4.2) | Crude | 1.16 (0.68-1.97) | 0.585 | 0.679 |
|  |  |  | IPTW | 0.99 (0.62-1.60) | 0.980 |  |  | IPTW | 1.23 (0.71-2.11) | 0.464 |  |

Data are presented as n (%) for the per-protocol population. \*The first secondary endpoint was assessed by the win ratio approach, in the hierarchical order of all-cause death, stroke, myocardial infarction, BARC type 3 bleeding, revascularization, and BARC type 2 bleeding. BARC, Bleeding Academic Research Consortium; CI, confidence interval; APT, dual antiplatelet therapy; HR, hazard ratio; IPTW, inverse probability of treatment weighting.

Notably, when assessed by the hierarchical composite of clinically relevant ischemic and bleeding events, stepwise de-escalation yielded significantly more wins relative to standard DAPT both in the multivessel disease (17.3% wins versus 12.0% wins; win ratio_IPTW_: 1.48, 95% CI: 1.02-2.16, P = 0.041) and single-vessel disease subgroups (12.8% wins versus 8.8% wins; win ratio_IPTW_: 1.47, 95% CI: 1.05-2.07, P = 0.025; P_interaction_ = 0.984; Table 2 and Figure 4).

**Figure 4.**
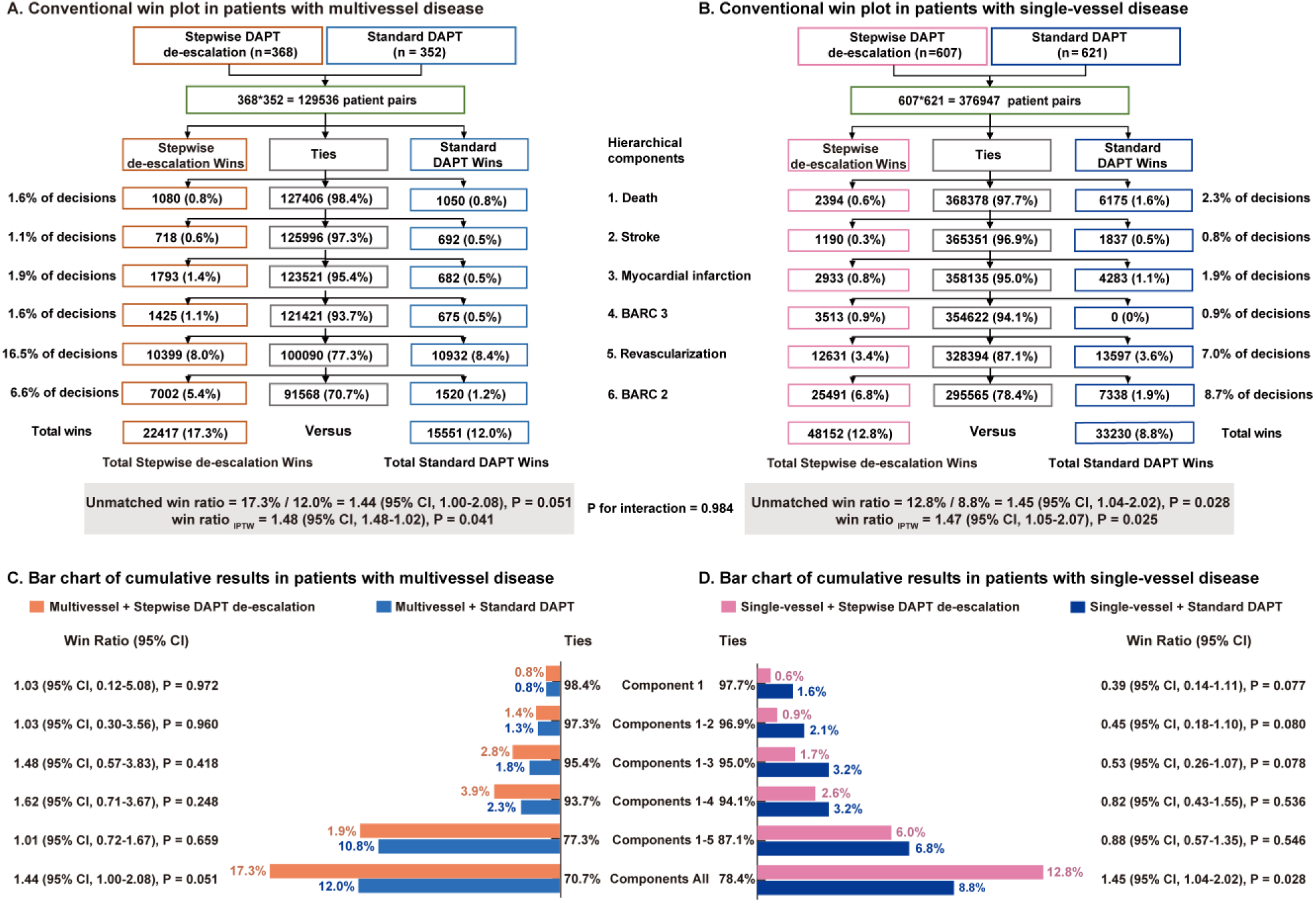
Win ratio diagram for the first secondary endpoint stratified by subgroups and treatment allocation. Conventional win plots among patients with multivessel disease **A** and single-vessel disease **B**, respectively. Bar charts showing cumulative results for patients with multivessel disease **C** and single-vessel disease **D**, respectively. BARC, Bleeding Academic Research Consortium; DAPT, dual antiplatelet therapy; IPTW, inverse probability of treatment weighting.

The results in the PP population were consistent with those in the ITT population (Figure S3, Tables S3 and S5). The clinical outcomes stratified by the number of diseased coronary vessels (1-vessel, 2-vessel, and 3-vessel disease) were presented in Figures S4 and S5. Cumulative event rates increased with rising coronary disease burden for NACE and PoCE (both P_trend_ < 0.001).

## DISCUSSION

In this prespecified analysis of the REC-CAGEFREE II trial, we found that:

1. Compared with patients with single-vessel disease, those with multivessel disease exhibited a higher prevalence of cardiovascular comorbidities and were associated with a significantly higher risk of NACE at 12 months.
2. No significant interaction was observed between vessel disease subgroups and treatment allocation for NACE. Compared with the standard DAPT group, the stepwise de-escalation group was associated with a lower risk of clinically relevant bleeding and no excess risk of ischemic events in both single-vessel and multivessel disease subgroups.
3. In the hierarchical analysis of clinically relevant ischemic and bleeding events, the stepwise de-escalation group was associated with greater net clinical benefit than standard DAPT in both subgroups.

Although dedicated randomized studies evaluating DCB strategies specifically for multivessel disease are still lacking, promising evidence from observational studies continues to accumulate. In a retrospective study comparing a DCB-based strategy with second-generation DES in patients with multivessel disease, the DCB approach significantly reduced stent burden during multivessel PCI and had a lower rate of major adverse cardiovascular events than DES-only treatment.^26^ Furthermore, in the prospective DCB-only All-Comers Registry of 1025 patients treated with paclitaxel-coated balloons (56.4% with multivessel disease), target lesion revascularization occurred in 3.2% of lesions, major adverse clinical events in 6.8%, and MI in 3.4% of patients at 9 months.^27^ Similarly, the EASTBOURNE prospective registry, which included nearly two-thirds of patients with multivessel disease, further demonstrated the safety and efficacy of sirolimus-coated balloons across a broad population with coronary artery disease.^28^ Highlighting the clinical relevance of these findings, recent randomized data from the REC-CAGEFREE II trial showed that multivessel disease accounted for 37% of the DCB-treated cohort, a proportion that closely aligns with the real-world prevalence (30% to 40% of the coronary disease population).^29^

Patients with multivessel disease had higher ischemic risk compared with single-vessel disease.^30–32^ This elevated risk profile is driven not only by anatomical and procedural complexity, but also by a heavy clustering of patient-level comorbidities, such as advanced age, CKD, diabetes mellitus, hypertension, and previous cardiovascular events.^33,34^ Consequently, patients with multivessel disease generally experience more complications, and together with multivessel disease itself, these factors contribute to a higher risk of adverse clinical outcomes. In the PEGASUS-TIMI54 trial, the multivessel disease cohort exhibited a higher prevalence of hyperlipidemia, peripheral artery disease, and prior revascularization compared with the non-multivessel disease cohort.^35^ These patterns were observed in other prior coronary artery disease studies.^2,3,36^ In line with these observations, patients with multivessel disease in our study presented with a significantly higher baseline comorbidity burden and a correspondingly elevated ischemic risk profile. Given the heightened ischemic vulnerability of this high-risk population, optimal DAPT duration and intensity are critical for balancing ischemic protection against bleeding safety.

Current guidelines default to 12-month DAPT for patients presenting with ACS after DES implantation.^37,38^ Anatomically, multivessel disease, with its high plaque burden and lesion complexity, intuitively supports extended antiplatelet therapy.^39^ Although dedicated evidence on DAPT regimens in ACS patients with multivessel disease treated with DCB is currently lacking, DES-era evidence shows that short DAPT followed by ticagrelor monotherapy consistently reduces bleeding without sacrificing ischemic safety.^14,16,17^ In contrast to DES, DCBs leave no permanent struts or polymers, eliminating persistent platelet activation and accelerating endothelial restoration. This provides a strong rationale for shortening DAPT after DCB angioplasty.^40^ In the large-scale, prospective EASTBOURNE registry, where patients with ACS accounted for 46.5% of the total cohort, a 6- to 12-month DAPT regimen was prescribed.^28^ However, the subsequent post hoc analysis comparing single antiplatelet therapy against DAPT demonstrated that single antiplatelet therapy following DCB angioplasty for both de novo and in-stent restenosis lesions was safe and effective, and could help to contain the risk of bleeding in a selected population.^41^ Another two-center real-world registry study, predominantly including patients with ACS and multivessel disease, reached the same conclusion: patients treated with a DCB-only strategy had a shorter DAPT duration than those receiving a hybrid strategy, and this strategy was associated with low lesion-level event rates and acceptable midterm clinical outcomes.^42^

In the current study, no significant interaction was observed between vessel disease subgroups and treatment allocation for the primary endpoint. Compared with the standard DAPT group, the stepwise de-escalation group was associated with a lower risk of clinically relevant bleeding without an increase in ischemic events in both single-vessel and multivessel disease subgroups. Conventionally, multivessel disease is positioned on the high ischemic risk side, an attribute that intuitively argues for maintaining, or even prolonging, rather than abbreviating, antiplatelet therapy.^5,6^ Our results indicated that the bleeding benefit of stepwise de-escalation was not achieved at the expense of ischemic safety even in this high-risk population. Furthermore, in the hierarchical analysis, the stepwise de-escalation group was associated with a greater net clinical benefit than standard DAPT in both subgroups. Our findings align with the DES-era trials, such as T-PASS and ULTIMATE-DAPT, in which abbreviated DAPT followed by ticagrelor monotherapy reduced bleeding while maintaining ischemic outcomes, with no significant interaction by multivessel disease.^16,17^ Taken together, these results provide evidence that, following DCB angioplasty, antiplatelet intensity can be reduced without compromising ischemic protection, even among patients with multivessel disease. Nevertheless, because subgroup analysis is inherently exploratory, these findings should be regarded as strictly hypothesis-generating and merit confirmation in dedicated trials for multivessel disease.

## LIMITATIONS

Our study has several limitations. First, as a subgroup analysis, this study was limited by reduced statistical power to detect true between-group differences and an increased risk of type I error arising from multiple testing; accordingly, these findings should be regarded as strictly hypothesis-generating. Second, randomization in the REC-CAGEFREE II trial was not stratified by vessel disease status; baseline imbalances between treatment groups may exist within subgroups. Although two statistical adjustment methods were employed, the potential for residual unmeasured confounding cannot be entirely excluded. Third, all patients were treated with paclitaxel-coated balloons only, because sirolimus-coated balloons were not commercially available in China during the study period. Therefore, the results cannot be extrapolated to sirolimus-coated balloons. Fourth, the present study was conducted solely in an East Asian population from China, and the generalizability of these results to other ethnicities requires further validation.

## CONCLUSIONS

Among patients with ACS undergoing DCB-only angioplasty, those with multivessel disease were associated with a higher risk of NACE than those with single-vessel disease. Stepwise DAPT de-escalation and standard DAPT exhibited similar risk-benefit profiles in both subgroups. Given the exploratory nature of the subgroup analysis, these findings should be regarded as strictly hypothesis-generating.

## Data Availability

The data that support the findings of this study are available from the corresponding author upon reasonable request.

## Acknowledgments

The authors thank the participants, their families, and all investigators involved in this study.

## Supplemental Material

Table S1-S5; Figure S1-S5

## ABBREVIATIONS

ACS: Acute coronary syndromes
DCB: Drug-coated balloon
DAPT: Dual antiplatelet therapy
NACE: Net adverse clinical events
PCI: Percutaneous coronary intervention
DES: Drug-eluting stent
MI: Myocardial infarction
BARC: Bleeding Academic Research Consortium

